# Cleaning and Re-Use of cobas® 6800/8800 Processing Plates for the SARS-CoV-2 Assay

**DOI:** 10.1101/2020.10.09.20209601

**Authors:** Matt Sperling, Ryan Relich, Bryan Schmitt, Drew Bell, Lauren Cooper

## Abstract

Real-time reverse transcription polymerase chain reaction (rRT-PCR) is the primary method used for the detection and diagnosis of infections caused by severe acute respiratory syndrome coronavirus 2 (SARS-CoV-2). Since SARS-CoV-2’s entrance into the United States, numerous clinical laboratories and *in vitro* diagnostic companies have developed rRT-PCR assays, some requiring specialized materials and reagents. One such assay includes the cobas® SARS-CoV-2 Qualitative Assay for use on the cobas® 6800/8800 (Roche Molecular Systems, Inc.). Since initiation of this assay at our facility, our ability to run testing at full capacity has been limited due to restricted supply of the omni cobas® Processing Plate (Product Number 05534917001), a 96 deep well plate used for all sample processing and total nucleic acid extraction via MagNA Pure magnetic beads. To work around this limiting factor, we have successfully designed and tested a cleaning protocol utilizing the widely available laboratory resources of bleach, ethyl-alcohol and autoclaving, for omni cobas® Processing Plate re-use.

## Introduction

Severe acute respiratory syndrome coronavirus 2 (SARS-CoV-2), the causative agent of coronavirus disease (COVID-19), is a positive single stranded RNA virus (+ ssRNA)^1^. Due to the nature of the virus’ nucleic acid, as well as the speed, sensitivity and specificity of molecular diagnostics, real-time reverse transcription polymerase chain reaction (rRT-PCR) is the primary method used for the detection and diagnosis of SARS-CoV-2. Beginning in March of 2020, numerous SARS-CoV-2 rapid tests received Emergency Use Authorization (EUA) from the U.S. Food and Drug Administration (FDA), including the cobas® SARS-CoV-2 Qualitative Assay for use on the cobas® 6800/8800 (Roche Molecular Systems, Inc.)^2,3^. Due to the overwhelming demand for testing reagents and consumables, governments, international agencies, health systems and *in vitro* diagnostic (IVD) companies, including Roche, have had to resort to resource allocation^4^.

At our institution, Indiana University Health (Indianapolis, IN, USA), one specific component of the cobas® SARS-CoV-2 Qualitative Assay was identified as our test capacity limiting factor, the omni cobas® Processing Plates (Product Number 05534917001). This processing plate is a 96 compartment, deep well plate used for sample processing and total nucleic acid extraction via MagNA Pure magnetic beads, specific for the cobas® 6800/8800. Several factors played into this product being a constraining factor: national medical grade plastic shortage resulting in strict allocation by Roche, continued need for the use of other assays on the cobas® 6800/8800 which also require the same processing plates, and cobas® automatic disposal and, therefore loss of processing plates in the event of run failure.

To work around this issue, we devised and tested a cleaning process for the re-use of omni cobas® Processing Plates. A few considerations were made in designing the cleaning process. First, since + ssRNA is directly infectious, safety was a primary focus. Second, we wanted to ensure that no residual viral nucleic acid remained in the processing plate after the cleaning protocol. Lastly, we wanted to assure that the cleaning process did not impair future use, including extraction of viral nucleic acid, or introduction of PCR inhibition from the processing plate. Grounded in these considerations, we have successfully designed and tested a cleaning protocol for omni cobas® Processing Plates that utilizes a BSII hood, bleach, ethyl-alcohol and autoclaving that allows for reuse with the SARS-CoV-2 assay.

## Methods

### Soaking of Processing Plates

Processing plates were retrieved from the waste bins onboard the cobas® 6800/8800 and brought under the BSII hood for processing. Plates were fully submerged in a 20% bleach solution in a nonreactive plastic storage container with strips of magnets (LOVIMAG, Product Number BAR8P) adhered to the side of the container to limit the movement of residual magnetic beads in the solution. Plates were soaked for at least 20 and up to30 minutes.

### Rinsing of Processing Plates

Next, a 10% bleach solution in a squeeze bottle was used to fill each well of the processing plate, and a transfer pipet was used to remove bleach solution and residual magnetic beads from the wells. This step was then repeated once. Lastly, a 10% bleach solution was placed into a non-reactive plastic container and the processing plates were fully submerged for two minutes. Next, a transfer pipette was used to remove any dried magnetic beads from the processing plate wells. Once complete, the processing plate was drained and placed upon a lint-free WypAll® (Kimberly Clark Worldwide, Inc.) towel until dry.

### Removal of Remnant Bleach

To remove any remnant bleach from the processing plates, 95% ethanol was used to fill the wells and cover the top of the processing plates. A transfer pipet was then used to remove all ethanol and plates were wrapped with a lint-free WypAll® (Kimberly Clark Worldwide, Inc.) towel and knocked 5 times on the tabletop to remove as much residual ethanol was possible. Plates were then left to air dry.

### Autoclaving

To shield from degradation during the autoclaving processes, processing plate barcodes were painted with clear nail polish and left to air dry. Next, a total of four processing plates were placed in a 12” x 18” autoclave pouch, insuring barcodes on the processing plates were facing inwards. Autoclave bags with processing plates were then autoclaved for 40 min at 121°C to both deactivate residual bleach and to sterilize the processing plates. Once the plates were removed from the autoclave and cooled to room temperature, any barcodes deemed to be too damaged for reading were replaced with re-printed barcodes affixed overtop with cellophane tape. Additionally, all plates were marked to denote the number of autoclave cycles it had encountered; for this study, a single processing plate was allowed three autoclave cycles before being discarded.

### Validation of Cleaning Processes

Four processing plates previously used to test patient samples for SARS-CoV-2 were cleaned by the above protocol. Hyclone™ (Cytiva Life Sciences, LLC.) molecular grade water samples were then run on the cleaned plates to determine if any residual nucleic acid remained after cleaning. For this assessment, we would expect all “water only” samples to be negative for SARS-CoV-2 specific targets (targets 1 and 2). Additionally, four positive patient samples and all sample internal control (IC) cycle threshold (Ct) values were evaluated for extraction inhibition due to the cleaning process. If extraction inhibition were to occur, we would expect significant Ct value increases in the SARS-CoV-2 specific targets for the positive patient samples, and significant increases in the IC Ct values for all sample types, including “water only”.

### Statistical Analysis

A two-sample t-test was used to calculate the *p*-value of IC Ct values before and after the above processing plate cleaning protocol.

## Results

SARS-CoV-2 detection in patient samples using four processing plates, prior to cleaning, can be found on **Table 1**. In total, 194 samples were tested over four unique processing plates where each plate had its own positive and negative control. Overall, 138 (74.19%) of patient samples were negative, 39 (20.97%) were positive, 8 (4.30%) were inconclusive (only target 2 was identified), and 1 (0.54%) was invalid (no internal control was identified)^3^.

**Table 1.** SARS-CoV-2 patient samples results prior to processing plate cleaning.

| <u>Total (n=194)</u> | <u>Sample Type</u> | <u>Target 1 Average Ct</u> | <u>Target 2 Average Ct</u> | <u>IC Average Ct</u> |
| --- | --- | --- | --- | --- |
| 4 | Positive control | 33.92 | 35.95 | 33.12 |
| 4 | Negative Control | - | - | 32.78 |
| 138 | Negative Patient | - | - | 33.61 |
| 39 | Positive Patient | 26.22 | 28.90 | 33.61 |
| 8 | Inconclusive | - | 32.43 | 33.31 |
| 1 | Invalid | - | - | - |

After performing the above cleaning protocol, the same four unique processing plates were run with positive and negative controls on each plate—all of which resulted as expected—and a total of 99 “water only” samples (**Table 2**). Ninety-eight (98.99%) “water only” samples resulted “negative” while a single sample (1.01%) resulted “invalid”. No significant differences in internal control Ct values were noted (*p*≤0.001). Additionally, using one of the four plates, four positive patient samples from Table 1 were rerun after the cleaning process and changes in the Ct values for SARS-CoV-2 specific were calculated (**Figure 1**).

**Table 2.** Post cleaning SARS-CoV-2 controls and “water only” samples.

| <u>Total (n=112)</u> | <u>Sample Type</u> | <u>Target 1 Average Ct</u> | <u>Target 2 Average Ct</u> | <u>IC Average Ct</u> |
| --- | --- | --- | --- | --- |
| 4 | Positive control | 33.62 | 35.68 | 33.35 |
| 4 | Negative Control | - | - | 33.09 |
| 98 | Water Only: Negative | - | - | 32.85 |
| 1 | Water Only: Invalid | - | - | - |

**Figure 1.**
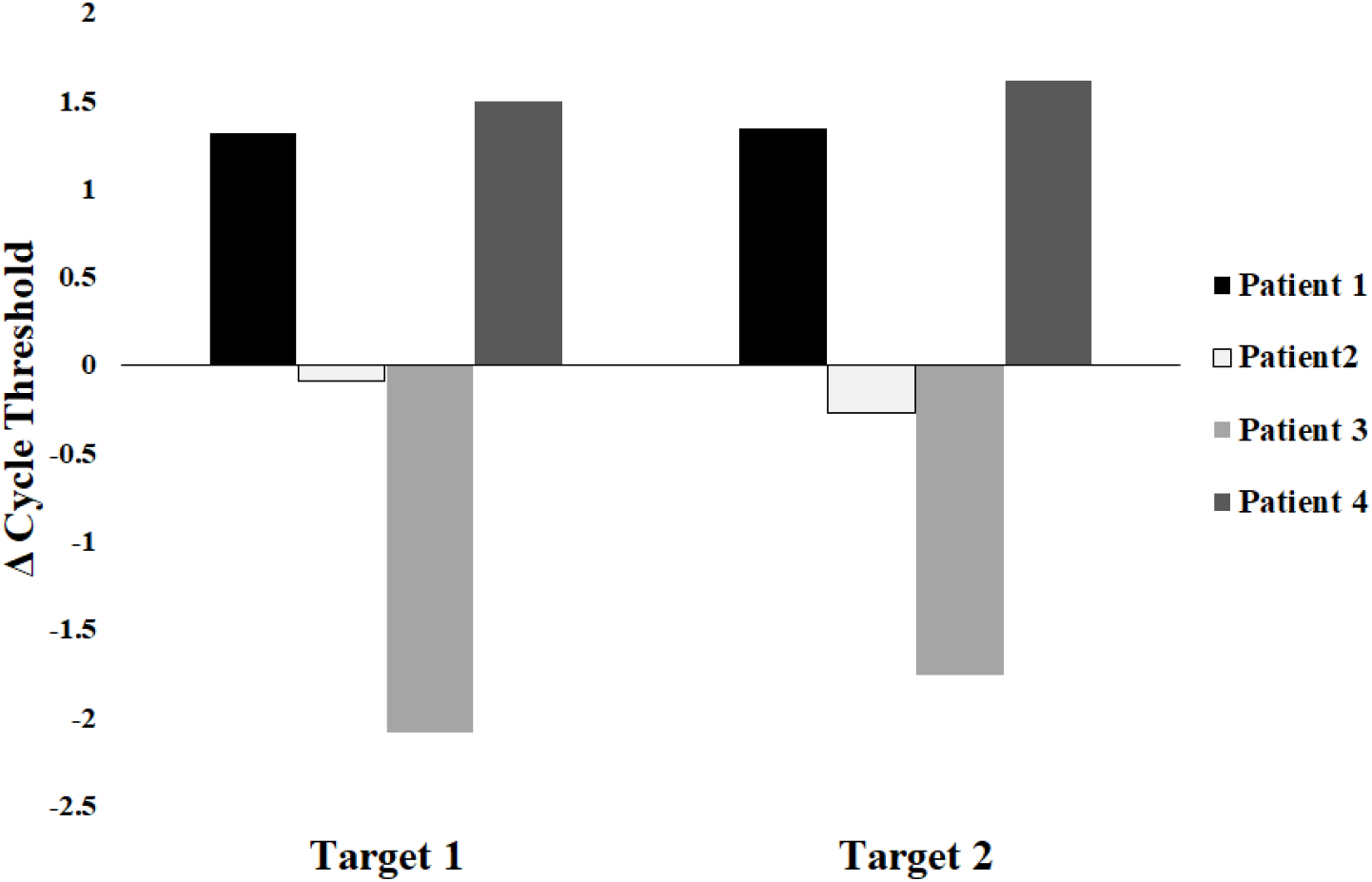
Changes in cycle threshold (Ct) values from positive patient samples before and after processing plate cleaning.

## Summary

The goal of this study was to design and test a cleaning protocol for omni cobas® Processing Plates that was safe, resulted in no residual viral nucleic acid carryover, and did not inhibit nucleic acid extraction. From these data, we show that four processing plates used in patient sample testing resulted in a mixture of positive (n=39), negative (138), inconclusive (8), and invalid (1) patient samples, along with positive and negative controls for each plate (**Table 1**). During the re-use of these same four plates, the “water only” samples (n=99) show that no measurable nucleic acid survived the above cleaning process (**Table 2**). In addition, no measurable changes occurred between the IC Ct values for samples from before and after cleaning (*p* ≤0.001). Lastly, we show that changes with SARS-CoV-2 specific targets from positive patient samples both before and after processing plate cleaning are within the normal range of precision variation^3^.

As CoVID-19 overwhelms healthcare systems around the world, hospitals, laboratories, collection centers are confronting shortages of personal protective equipment (PPE), test kits, ventilators, and more. To meet these challenges, as health care providers, we must prioritize conserving, substituting, adapting, re-using and re-allocating what resources we do have. Examples of meeting challenges has been displayed in many ways over the last few months including: using a single swab for both NP and oropharyngeal specimen collection, institutions preparing their own viral or universal transport media, 3D printing of nasopharyngeal swabs, ventilator parts, and various PPE, and the re-use of single-use, disposable PPE^5–7^.

However, it should be stated that even when laboratory supplies reach a “non-critical” level, it should be a priority of all laboratories to reduce its dependence on single-use plastics. In 2015, a study was done that suggests research laboratories alone produces over 5.5 million metric tons of plastic waste a year^8^. Although finding ways to conserve, substitute and re-use biohazard laboratory waste can be difficult, there are several ways that working with commercial suppliers can impact how much waste a laboratory produces. First, suppliers could allow for more bulk orders to decrease the need for excess packaging, authorize packaging to be sent back for re-use, supply laboratories with cleaning protocols for re-use of “single-use” items, and allow for the return of “single-use” items for recycling. Additionally, laboratories could find additional uses for leftover packaging, conserve plastic use where able, switch to glass containers and tools, and develop their own protocols for cleaning “single-use” items.

## Data Availability

All data presented in this manuscript is available by request to the corresponding author.

## Notes

### Competing Interest Statement

The authors have declared no competing interest.

### Author Declarations

IRB Indiana University School of Medicine

